# External validation of dynamic clinical states in acute stroke: transportability and independent rediscovery across 176 hospitals

**DOI:** 10.64898/2026.09.07.26362407

**Authors:** Ping Lei, Yanyi Xu, Yuxia Zhang

**Author notes:** **Corresponding author:** Ping Lei, Institute of Global Health, University of Geneva, Geneva, Switzerland.

## Abstract

**Objective:** Four dynamic clinical states were identified in the first 72 h of stroke intensive care in one centre. We tested whether this representation transports to an independent multicentre cohort, predicts the next state, and is independently recovered.

**Materials and Methods:** 8279 adults in 176 hospitals (eICU-CRD v2.0) contributed 70 630 six-hour windows. Phenotype, dictionaries and eligibility rules were frozen before any state was assigned; the model was applied unchanged and judged against five prespecified criteria. Generalised estimating equations related state to subsequent organ support and ICU death; prediction was assessed against a persistence null at 6, 12 and 24 h; a model was fitted de novo in the strictest scope.

**Results:** All five transportability criteria were met in all four scopes. All nine state–outcome comparisons reproduced the direction of association, with exact rank order for invasive ventilation and ICU death; between-hospital intraclass correlations were 0.012–0.027. States persisted across 90.7% of pairs, so prediction was scored on change: AUROC (95% CI) 0.730 (0.723–0.736) externally, 0.735 (0.721–0.749) internally. Fitted de novo, eICU favoured four by BIC but three by restart reproducibility; neither recovered neurological impairment–low support, the other three matched closely in both (r = 0.962–0.991).

**Discussion:** The representation transports and carries forward-looking information but is not fully rediscoverable. The unrecovered state is defined by a combination, not a feature: eICU places its windows consistently but never pairs impairment with absent organ support.

**Conclusion:** Three of four states are strongly supported. Transportability and independent rediscovery are distinct and should be reported separately.

## Background and Significance

The early intensive care course of acute stroke is not static. Patients arrive with a severity that is documented once and then move: neurological responsiveness fluctuates, organ support is started and withdrawn, renal and respiratory function diverge from the admission picture within hours. A single admission phenotype cannot represent that movement, and the alternative — treating each six-hour window as an independent observation — discards the sequence that makes the course interpretable.

We previously described the first 72 h of intensive care after acute stroke in MIMIC-IV as movement among four dynamic states, recovered by a hidden Markov model over 21 routinely recorded variables.[1] About 40% of patients changed state during the observation period, and the current state was associated with subsequent organ support and with death after adjustment for baseline characteristics. Those results came from one academic centre. Internal reproducibility across admission eras is reassuring but is not external validation, and a latent structure recovered from a single institution’s records may encode that institution’s documentation habits as readily as it encodes clinical physiology.

Validation of a latent-state model is usually reported as a single verdict, and that conflates two questions with different answers. The first is whether the model transports: applied unchanged to new data, does it assign states that behave as the original states did? The second is whether the structure is independently rediscoverable: given the same variables and no knowledge of the original solution, would an analyst working in the new data arrive at it? A model can be entirely usable without being the structure the new data would select on its own. The distinction is not academic: the first question governs whether the representation can be applied, the second whether it describes anything outside the cohort that produced it.

A third property has received less attention still. A partition of windows that merely describes the data well need not carry any information about which patients are about to move. If a state representation is capturing clinical trajectory rather than documentation pattern, then the state a patient occupies now should say something about the state they will occupy next — and that predictive content, if it is real, should survive transport as the state definitions do.

Three questions therefore stand apart: whether the representation travels, whether it carries information about what comes next, and whether an analyst starting from the new data alone would have found it.

This study puts all three to an independent multicentre cohort. eICU-CRD[2] draws on 176 hospitals distinct from the discovery centre in geography, size, teaching status and electronic record system. We first apply the frozen discovery model there against prespecified success criteria, and test whether the transported states retain their outcome associations and their stability across hospitals. We then ask whether the state a patient occupies carries information about the state they occupy next, at horizons up to a day. Finally we set the discovery model aside and fit a model de novo, to ask what structure eICU supports on its own terms.

Reporting the transport result and the rediscovery result separately, rather than either one as a verdict, is the methodological contribution we intend this study to make.

## Materials and Methods

### Design, data and analysis order

Retrospective multicentre cohort study using eICU-CRD v2.0 as an external validation dataset for a state representation derived in MIMIC-IV v3.1.[3] Both databases are held at PhysioNet under credentialed access.[4] The analysis was ordered so that no step could be informed by a later one: cohort construction and variable harmonization were completed and archived before any state was assigned, and the frozen-model analysis was completed and archived before any model was fitted within eICU.

### Cohort and phenotype

Stroke ascertainment does not transport. In eICU, icd9code is a deterministic re-encoding of the free-text diagnosisstring, so the discovery study’s ICD list carries no independent information here. The phenotype was therefore rebuilt as an enumerated list of 35 diagnosisstring paths, clinically adjudicated, assigned to subtype, and frozen before extraction (Supplementary Table S1); the APACHE admission diagnosis was retained as an independent cross-check. Adults with a qualifying path and an ICU stay of at least 12 h were included, the first stay only, and observation ran to 72 h in twelve consecutive six-hour windows.

### Variable harmonization and hospital eligibility

All 21 model variables were mapped to eICU source tables, fields, units and aggregation rules, and window-level availability was computed identically in both databases before any forward fill (Supplementary Table S2). Two variables constrained the analysis. Glasgow Coma Scale subscores are recorded in 53.5% of eICU windows against 94.9% in discovery, and coverage is a hospital property: 36 of 176 hospitals chart them in fewer than half of windows. Invasive ventilation cannot be separated from non-invasive support in every hospital, and no hospital resolves it for more than 67% of its stroke patients, so ventilation is reported as a bracket rather than a point estimate. Because restricting to patients in whom these are determinable would enrich for the ventilated, both were resolved at hospital level: two prespecified eligibility rules define a GCS-eligible and a ventilation-ascertainable subset, and every analysis is reported in four scopes — the full cohort, each rule alone, and their intersection.

### Frozen-model transport

The discovery model, a hidden Markov model fitted with pomegranate,[5] was applied as a read-only object. Emission and transition parameters, imputation constants, scaling constants and state labels were frozen and exported before use; nothing was estimated from eICU during state assignment. An epsilon floor of 1e-4 was applied to the categorical emissions, verified not to alter discovery decoding. Five criteria were fixed in advance: per-state profile correlation ≥0.80, minimum state prevalence ≥5% of windows, transition rank correlation ≥0.80, maximum self-transition deviation ≤0.10, and an observed minimum profile correlation above the 95th percentile of a permutation null. Because the four organ-support variables are the least transportable elements of the feature set, a 17-variable treatment-free model was transported alongside as a prespecified specificity control.

### Outcomes, heterogeneity and independent refitting

Generalised estimating equations[6] with an exchangeable working correlation clustered by patient related the current state to new invasive ventilation and new vasoactive support within 12 h and to ICU death, adjusted for age, sex and stroke subtype. Between-hospital variation in state prevalence was estimated with random-intercept linear probability models across hospitals meeting both eligibility rules. A hidden Markov model was then fitted de novo within eICU, in the strictest scope so that the comparison rests only on hospitals where both measurements are dependable, across a range of state counts, selected on Bayesian information criterion[7] and on stability across random restarts, measured by the adjusted Rand index,[8] and compared with the discovery states by profile correlation.

### Prediction of the next state

The state process is persistent, so overall accuracy is uninformative: a rule predicting no change scores about 0.91 while identifying nobody. Prediction was therefore judged by discrimination for the binary event that the state differs in the following window, for which that rule is uninformative by construction. Six predictors were arranged as a ladder, each rung permitted more information than the one below it: nothing at all (persistence); the decoded current state (the frozen transition matrix); the full observation history through the current window (the frozen model, forward filtered); the 21 variables recorded in the current window (multinomial logistic regression, then gradient boosting[9] without a linearity assumption); and the raw observation sequence (a gated recurrent network,[10] included as an empirical benchmark for a model not restricted to one window at a time). The first three rungs involve no fitting. The remainder were fitted on the MIMIC-IV training split alone and frozen before eICU was decoded. Horizons of 6, 12 and 24 h were examined, with the transition matrix raised to the corresponding power. Intervals are patient-clustered bootstrap percentiles, and comparisons between predictors are paired within the same resample.

### Prespecification

The protocol was written before extraction and amended fifteen times during it, each amendment dated, classified and archived with what was visible at the time (Supplementary Table S3). Fourteen were made with no outcome result visible in either database. The fifteenth added the prediction analysis after the preceding results were known; it altered no frozen parameter and its metric was fixed before any prediction result was computed, but it is post-hoc and is reported as exploratory.

## Results

### Cohort

The external cohort comprised 8279 adults with acute stroke in 176 hospitals, contributing 70 630 six-hour windows (Table 1). It is larger than the discovery cohort of 6368 patients. Median age was 68 years [IQR 57–79], against 69 in discovery; 48.5% were women; median ICU stay was 2.02 days [1.19–4.11]; ICU mortality was 7.3%.

**Table 1.** Characteristics of the eICU external validation cohort, overall and by stroke subtype. Subtype precedence follows the discovery study; stays carrying only the unspecified stroke path are reported as a separate column and their exclusion is a prespecified sensitivity analysis.

| Characteristic | All patients | AIS | ICH | SAH | ICH+SAH | unspecified |
| --- | --- | --- | --- | --- | --- | --- |
| Patients, n | 8,279 | 3,198 | 1,736 | 548 | 104 | 2,693 |
| Age, median [IQR] | 68 [57–79] | 70 [59–80] | 68 [56–79] | 58 [48–69] | 62 [51–71] | 70 [59–79] |
| Female, n (%) | 4,015 (48.5) | 1,546 (48.3) | 786 (45.3) | 308 (56.2) | 58 (55.8) | 1,317 (48.9) |
| Neuro ICU, n (%) | 2,884 (34.8) | 1,042 (32.6) | 771 (44.4) | 293 (53.5) | 46 (44.2) | 732 (27.2) |
| Teaching hospital, n (%) | 2,829 (34.2) | 1,175 (36.7) | 674 (38.8) | 256 (46.7) | 35 (33.7) | 689 (25.6) |
| ICU stay, days, median [IQR] | 2.02 [1.19–4.11] | 1.92 [1.17–3.58] | 2.56 [1.45–5.18] | 4.90 [1.87–11.75] | 7.53 [2.45–12.89] | 1.84 [1.08–3.31] |
| 6-hour windows contributed, median | 9 | 8 | 11 | 12 | 12 | 8 |
| ICU mortality, n (%) | 601 (7.3) | 169 (5.3) | 204 (11.8) | 59 (10.8) | 17 (16.3) | 152 (5.6) |
| Hospital mortality, n (%) | 1,091 (13.2) | 365 (11.4) | 337 (19.4) | 88 (16.1) | 26 (25.0) | 275 (10.2) |

Two-thirds of patients left the ICU or died within 72 h. For most of this cohort the observation period is therefore not an early fragment of the ICU course but the whole of it.

### The frozen model transports

Applied unchanged, the discovery model met all five prespecified criteria in all four analysis scopes (Table 2). The strictest scope — hospitals satisfying both eligibility rules — is the one to read. There the weakest per-state profile correlation was 0.912 and the transition rank correlation 0.974, and 38.1% of patients changed state at least once, against 40.3% in discovery.

**Table 2.**
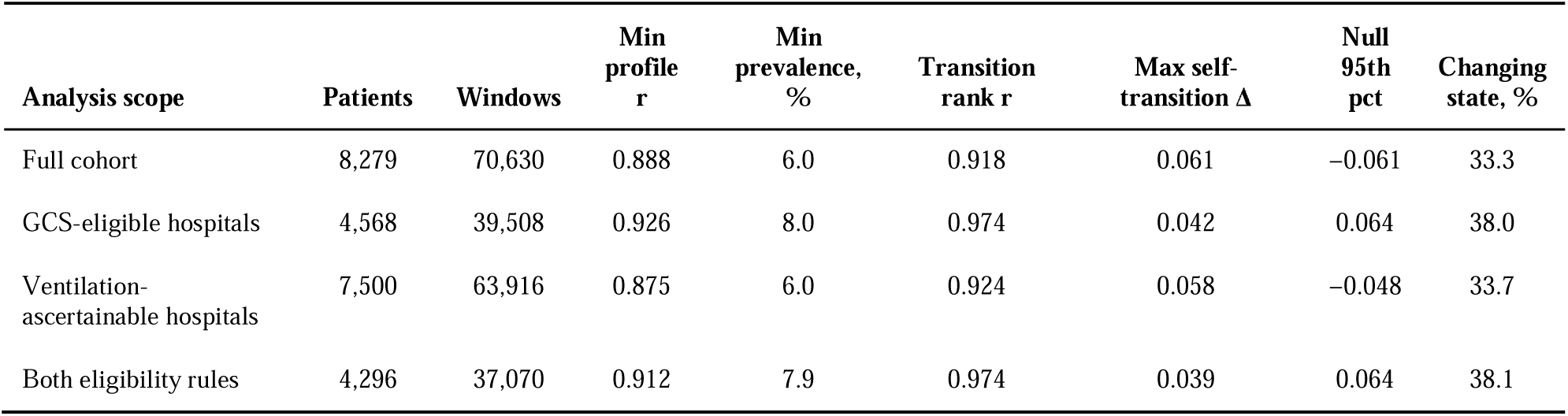
Frozen-model transport against the five prespecified criteria. Criterion thresholds: per-state profile correlation ≥0.80; every state ≥5% of windows; transition rank correlation ≥0.80; maximum self-transition deviation ≤0.10; observed minimum profile correlation above the 95th percentile of the permutation null. All five are met in all four scopes. The proportion of patients changing state is descriptive and is not a pass/fail criterion; the discovery value is 40.3%.

| Analysis scope | Patients | Windows | Min<br>profile<br>r | Min<br>prevalence,<br>% | Transition<br>rank r | Max self-transition $\Delta$ | Null<br>95th<br>pct | Changing<br>state, % |
| --- | --- | --- | --- | --- | --- | --- | --- | --- |
| Full cohort | 8,279 | 70,630 | 0.888 | 6.0 | 0.918 | 0.061 | −0.061 | 33.3 |
| GCS-eligible hospitals | 4,568 | 39,508 | 0.926 | 8.0 | 0.974 | 0.042 | 0.064 | 38.0 |
| Ventilation-ascertainable hospitals | 7,500 | 63,916 | 0.875 | 6.0 | 0.924 | 0.058 | −0.048 | 33.7 |
| Both eligibility rules | 4,296 | 37,070 | 0.912 | 7.9 | 0.974 | 0.039 | 0.064 | 38.1 |

Two prespecified controls test the obvious objections.

The first is that transport reflects how each hospital records treatment rather than the patients’ physiology. Comparing the 17 continuous variables alone, with the four organ-support variables removed, left the weakest correlation at 0.913 — unchanged.

The second is that transport is an artefact of imputing the missing motor subscore. Restricting both databases to windows where it was actually measured raised the weakest correlation to 0.938. Imputation was attenuating transport, not creating it.

### The states retain their outcome associations

No outcome information was used at any point in state assignment. Nevertheless all nine state-by-outcome comparisons reproduced the direction of association observed in discovery, with complete ordinal preservation for new invasive ventilation and for ICU death (Table 3). For new vasoactive support the two most severe states exchanged rank between databases (renal dysfunction 4.91 and respiratory support 5.00 in discovery, against 3.60 and 3.29 here), a reversal within a gap of 0.09 in the discovery estimates. Relative to the neurologically preserved state, the odds of new invasive ventilation within 12 h rose from 2.01 (95% CI 1.58–2.57) in neurological impairment–low support to 7.52 (6.12–9.25) in the respiratory-support state, and the discrete-time odds of ICU death from 9.91 (6.63–14.82) in renal dysfunction to 44.51 (31.38–63.13) in respiratory support. One mortality estimate rested on two events in each database and was suppressed under the prespecified rule.

**Table 3.** Adjusted odds ratios for the current state and subsequent events, reference state neurologically preserved–low support. Generalized estimating equations with an exchangeable working correlation clustered by patient, adjusted for age, sex and stroke subtype. † Suppressed under the prespecified rule of fewer than ten events; the discovery study’s estimate for the same state also rested on two events. All nine comparisons reproduce the direction of effect.

| Outcome | State | MIMIC-IV OR (95% CI) | eICU OR (95% CI) | eICU events |
| --- | --- | --- | --- | --- |
| New invasive ventilation within 12 h | Neurological impairment–low support | 1.64 (1.14–2.35) | 2.01 (1.58–2.57) | 149 |
| New invasive ventilation within 12 h | Neurological impairment–renal dysfunction | 3.25 (2.32–4.55) | 4.46 (3.68–5.40) | 408 |
| New invasive ventilation within 12 h | Neurological impairment–respiratory support | 4.28 (3.10–5.92) | 7.52 (6.12–9.25) | 296 |
| New vasoactive support within 12 h | Neurological impairment–low support | 1.21 (0.81–1.79) | 1.69 (1.17–2.44) | 53 |
| New vasoactive support within 12 h | Neurological impairment–renal dysfunction | 4.91 (3.79–6.36) | 3.60 (2.82–4.59) | 175 |
| New vasoactive support within 12 h | Neurological impairment–respiratory support | 5.00 (4.07–6.13) | 3.29 (2.52–4.28) | 134 |
| ICU death, discrete-time hazard | Neurological impairment–low support | 0.21 (0.05–0.87) | 0.58 (0.14–2.40)† | 2 |
| ICU death, discrete-time hazard | Neurological impairment–renal dysfunction | 3.73 (2.48–5.59) | 9.91 (6.63–14.82) | 60 |
| ICU death, discrete-time hazard | Neurological impairment–respiratory support | 11.83 (9.00–15.55) | 44.51 (31.38–63.13) | 216 |

### Prevalence is stable across hospitals

Comparing hospitals to each other requires both measurements to be dependable at each one, so this analysis alone is restricted to the 38 of the 176 hospitals that satisfy both eligibility rules; the other analyses use the full cohort.

Across those 38 hospitals, intraclass correlations for state occupancy ranged from 0.012 to 0.027, indicating that only a small proportion of overall variation was attributable to between-hospital differences. Between-hospital standard deviations were 2.9 to 7.6 percentage points. Hospital-level prevalence of the largest state nonetheless spanned 46.6% to 86.3%, so low clustering is not the same as uniformity.

### The states carry information about the next window

The state process is persistent: 90.7% of eICU window pairs and 91.4% of held-out MIMIC-IV pairs occupy the same state six hours later. Every predictor examined therefore scores between 0.889 and 0.906 on overall accuracy, including the rule that predicts no change, and accuracy is reported only for completeness.

Judged on discrimination for change, the frozen model separates the windows that were about to change from those that were not, with no fitting of any kind (Table 4). The transition matrix alone, applied to the decoded current state, reached an area under the ROC curve of 0.684 (95% CI 0.679–0.691); propagating the filtered posterior over the full observation history raised this to 0.730 (0.723–0.736), against 0.735 (0.721–0.749) in the MIMIC-IV test split. The external AUROC was therefore 0.005 lower than the internal value. Predictors fitted on MIMIC-IV and frozen before eICU was decoded added 0.036 (0.030–0.041), and a recurrent network reading the raw observation sequence reached 0.787.

**Table 4.**
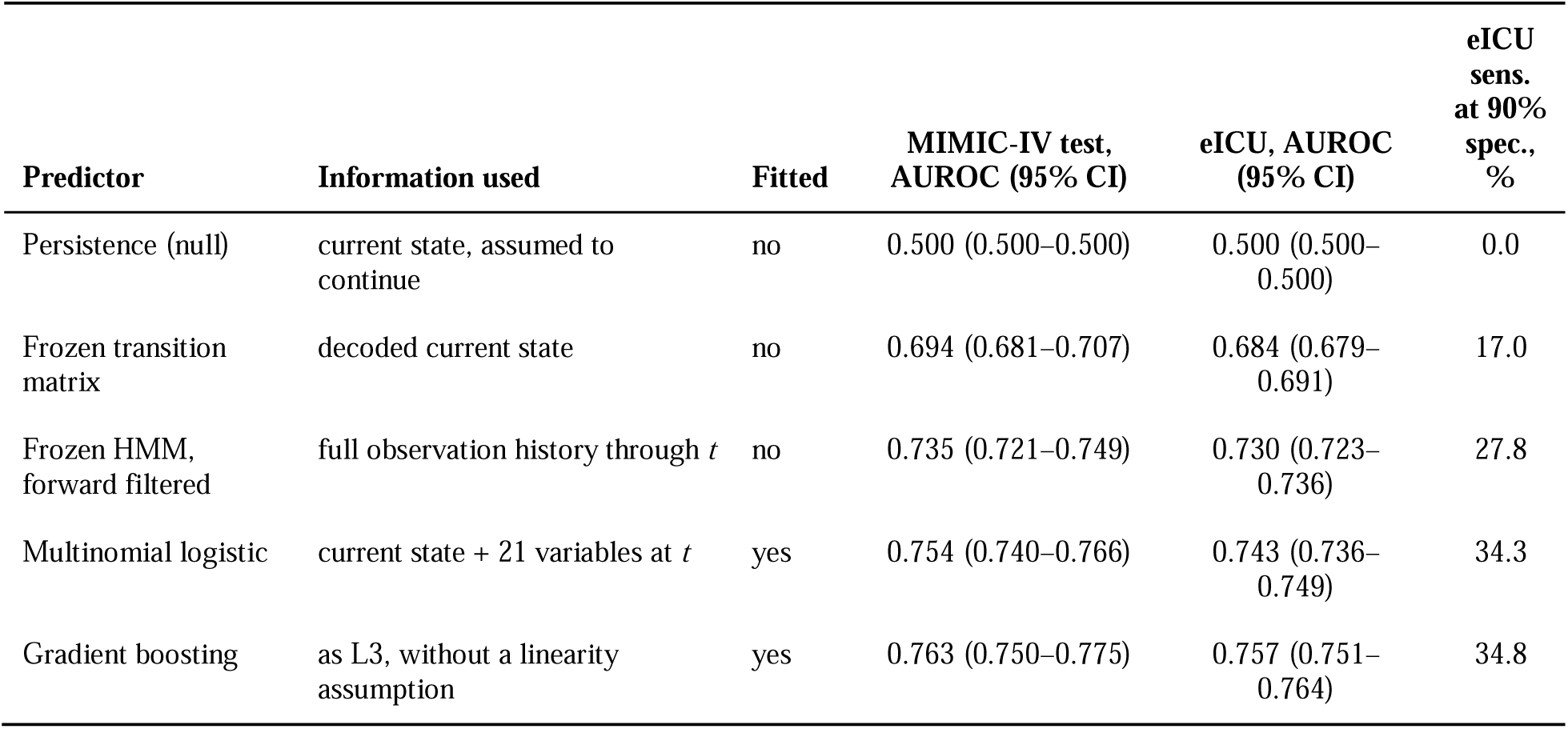
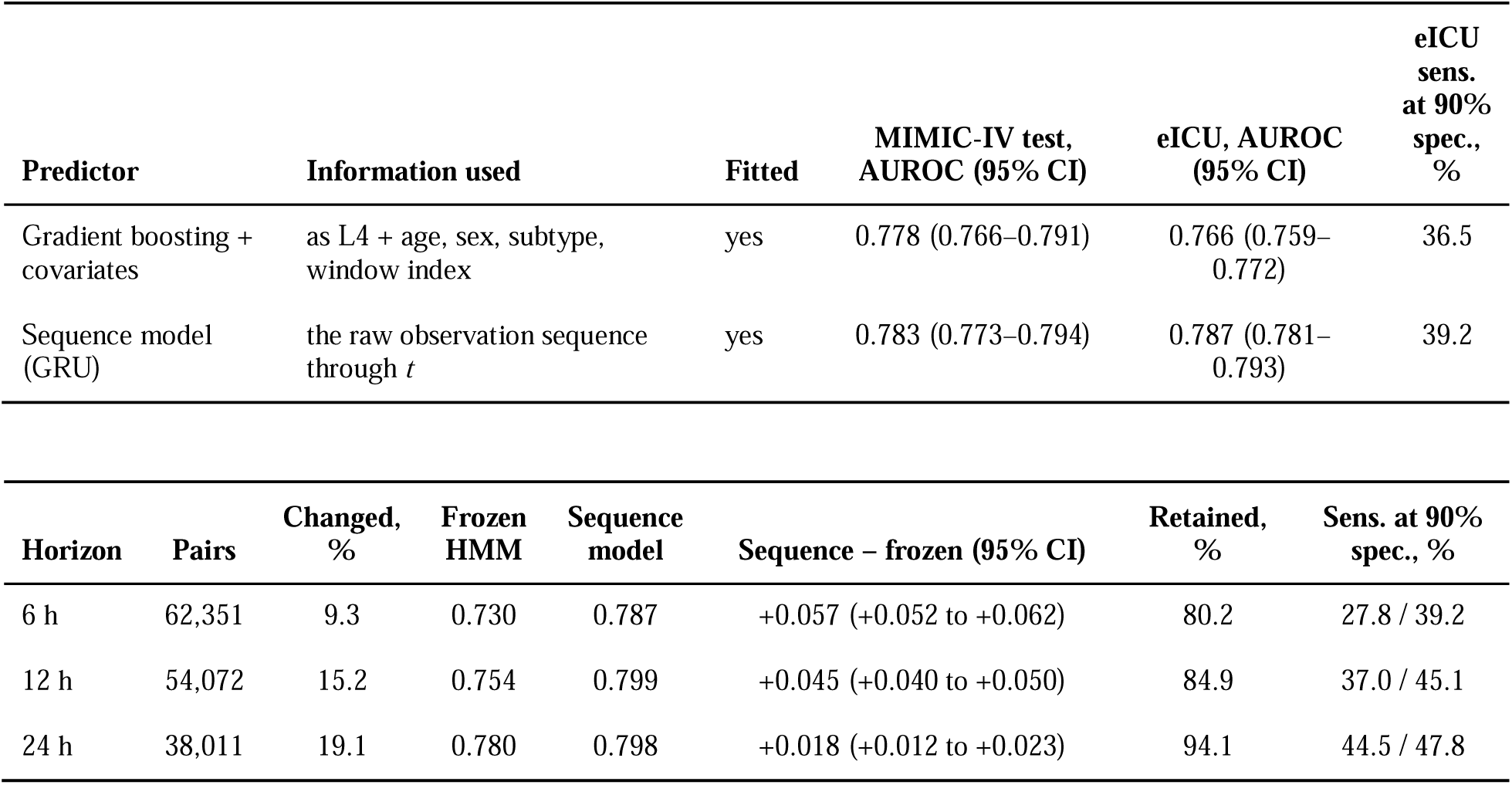
Prediction of the next state. The event is that the state differs in the following window; it occurs in 9.3% of eICU pairs, so a rule predicting no change scores 0.906 accuracy while identifying nobody, and discrimination for change is used instead. *Panel A, a 6-hour horizon:* predictors ordered by the information each is permitted to use. The first three involve no fitting and are the discovery model used for a different operation; the rest were fitted on the MIMIC-IV training split and frozen before eICU was decoded. The sequence model is included as an empirical benchmark and is the mean of three seeds. *Panel B, eICU across horizons:* the frozen transition matrix raised to the corresponding power; the fitted predictors were trained separately in the MIMIC-IV training split for each horizon and frozen before external evaluation, never refitted in eICU. Discrimination rises rather than decays, and the proportion retained — the discrimination above chance reached with no fitting, as a percentage of the sequence model’s, a fraction of discrimination and not of information — rises with it, so what the four states discard is concentrated at short range. The final column gives sensitivity at the threshold yielding 90% specificity, as frozen model / sequence model. It is reported because area under the curve summarises every threshold at once whereas a clinician uses one: the frozen model identifies 27.8% of transitions at six hours and 44.5% at twenty-four, so the longer horizons are more usable and not merely easier to rank, and none of the three reaches a sensitivity that would support alerting on an individual patient. Intervals are patient-clustered bootstrap percentiles; the horizon contrasts are paired within the same resample. Full results across all scopes are in Supplementary Table S4.

Discrimination does not decay as the horizon lengthens; it rises, and the frozen model rises much faster than the sequence model. Between 6 and 24 h the frozen representation improved from 0.730 to 0.780 while the sequence model improved from 0.787 to 0.798, so the gap between them narrowed from 0.057 to 0.018. Expressed as a fraction of the discrimination the sequence model achieves above the null, the frozen model supplied 80% at six hours and 94% at twenty-four. Holding the population fixed at the 24 h eligible set left the rise intact and slightly steeper, so it is not an artefact of the shrinking eligible set. Most of the discrimination the four states forgo is therefore concentrated at the shorter horizon: the representation tracks trajectory well and fluctuation less well. None of this constitutes a usable alarm. Held to a threshold silent for nine in ten stable windows, the frozen model identified 27.8% of the windows that changed at six hours, 37.0% at twelve and 44.5% at twenty-four, against 39.2%, 45.1% and 47.8% for the sequence model.

### Independent refitting did not recover all four discovery states

Fitted de novo in the strictest scope — the 4296 patients in the 38 hospitals satisfying both eligibility rules — eICU did not identify a uniquely preferred number of states. The four-state solution improved Bayesian information criterion over the three-state solution (754 215 against 826 358), whereas reproducibility across random restarts strongly favoured three (adjusted Rand index 0.991 against 0.840); five- and six-state models failed to converge. The two criteria disagree, and we report both rather than selecting between them, because the finding does not depend on the choice.

In the three-state solution each state corresponded closely to a discovery state: preserved–low support (r = 0.988), renal dysfunction (0.991) and respiratory support (0.971). The four-state solution matched the same three and spent its fourth on subdividing the preserved state, whose two components correlated 0.962 and 0.680 with it. Neither solution contained a state corresponding to neurological impairment–low support, whose highest correlation with any de novo state was 0.183.

## Discussion

### Transportability and independent rediscovery are different properties

“Does it validate?” is two questions, not one.

The first is whether the model *transports*: applied unchanged to new data, does it assign states that behave as the original states did? The second is whether the structure is *independently rediscoverable*: given the same variables and no knowledge of the original solution, would an analyst in the new data arrive at it?

Here the two questions gave different answers. Applied unchanged to 176 hospitals that contributed nothing to its construction, the discovery model met every prespecified criterion, reproduced the direction of association in all nine state-outcome comparisons, and carried information about a patient’s next state; across the 38 hospitals where both measurements are dependable, between-hospital clustering was slight. Refitted from scratch within those same 38 hospitals, the data did not reproduce that structure. Fit and reproducibility criteria disagreed on how many states eICU supports — four by Bayesian information criterion, three by stability across restarts — but the disagreement does not affect the conclusion: neither solution contained a counterpart to the fourth discovery state.

Both results are true, and one verdict cannot hold both. Reporting only the first, which is the usual practice, would have licensed the statement that all four states passed external validation. A reader would then take the fourth state to be as well founded as the other three. It is not.

We would encourage reporting the two separately, because they matter to different readers. Transportability is what matters to anyone who wants to apply the representation. Structural reproducibility is what matters to anyone who wants to read the states as features of the clinical course rather than as one defensible way of dividing it. The two failure modes also call for different responses: the first can be addressed by recalibration, the second cannot be resolved by recalibration alone — it is information about what the representation is.

A third lesson runs underneath both. Transporting a latent representation requires validating the measurement process as well as the model. Identical variable names did not imply identical measurement here, and the three obstacles we met between these two databases are documented in the Methods for others who will meet them.

### One state, not one database

The four-state framework did not fail; one state did. Three states are defined by a positive feature — preserved responsiveness, renal dysfunction, respiratory support — and all three were consistently supported across the analyses: they transport, they retain outcome associations, they are stable across hospitals, and they are independently rediscovered in both de novo solutions, with correlations of 0.971–0.991 at three states and 0.962–0.979 at four. The fourth, neurological impairment–low support, is defined by a combination rather than by a feature: the patient is impaired but is not receiving organ support. It is the smallest state, it contributed too few deaths for an odds ratio to be estimated, and it was the least well recovered when organ-support variables were removed.

Following its windows into the de novo solutions shows why it is not recovered, and the answer is not that it carries no signal (Supplementary Table S5). Two thirds of its windows are placed with the respiratory-support state under both the three-state and the four-state solution — 64.1% and 66.6% — and only 13.0% and 20.6% with the preserved state. They are assigned consistently along the neurological axis, not scattered. What eICU does not produce is the combination: no de novo state pairs neurological impairment with an absence of organ support. Given a fourth state to spend, eICU subdivides the largest state along urine output and metabolic variables instead, none of them neurological.

The state therefore appears to sit across two boundaries — grouped with the impaired states on the neurological axis, and with the unsupported states on the organ-support axis — and an algorithm partitioning along dominant axes assigns it to one side rather than isolating the intersection. Whether that reflects a property of this state or of the fitting procedure cannot be settled with one external database. We would report this as a four-state framework of which three are strongly supported and one requires reconsideration, and would suggest that clinical interpretation rest on the three.

### The representation carries predictive structure, and its losses are specific

A partition that merely described the data well would not, of itself, say anything about which patients are about to move. This one does, and it does so in hospitals that contributed nothing to its construction, with essentially no attenuation: 0.730 externally against 0.735 internally. The closeness of the internal and external values is further evidence that the predictive structure is not specific to the discovery institution.

Framing that comparison correctly matters, because the obvious summary is misleading here. Nine in ten windows are followed by a window in the same state, so a rule that predicts no change scores 0.91 accuracy while flagging nobody. We would encourage studies of persistent state processes to report the persistence rate alongside any accuracy they quote.

The four states are nonetheless a lossy summary, and the loss is specific. A recurrent network reading the raw sequence discriminates better at every horizon, so reducing a window to one of four labels does discard relevant information. But the loss is concentrated at short range: the frozen representation reaches 80% of the sequence model at six hours and 94% at twenty-four. Many six-hour transitions may represent brief excursions across a state boundary, whereas changes persisting to twenty-four hours are more likely to reflect sustained trajectories. The representation is an efficient summary of trajectory and a poor summary of fluctuation, which is the useful trade. It also indicates the incremental discrimination achievable through more flexible modelling of the same 21 variables: the sequence model gains 0.018 at twenty-four hours.

Substantially better day-scale prediction may therefore require information the present feature set does not contain — imaging, stroke-specific severity measures, or treatment decisions — rather than more flexible models applied to the same variables.

### Limitations

Invasive ventilation is bracketed rather than measured, and its lower bound cannot be compared with the discovery value at face value. GCS availability is roughly half that of discovery; the eligibility rule removes the hospital-level confound but the full-cohort analysis still carries it. eICU contains no post-discharge survival, and comorbidity could not be summarised comparably, so models adjust for a reduced covariate set. Five- and six-state models failed to converge in both databases, so a richer structure is untested. eICU covers 2014–2015 and predominantly community hospitals, so era and case-mix differ; reproduction of prognostic ordering across that difference is a strength, but absolute rates are not comparable.

Three limitations attach to the prediction analysis. It was added after the preceding results were known and is exploratory, although no frozen parameter was altered and its metric was fixed before any prediction result was computed. The sequence-model benchmark is empirical: a different architecture could achieve higher discrimination and therefore lower the proportion retained by the frozen representation. And the predicted target is a state assigned by the frozen model, decoded over the complete stay, so a network trained on those labels can learn to approximate that decoding from past data alone; the margin between it and the frozen model should therefore be read as an empirical upper bound on the loss in discrimination associated with the four-state representation, rather than as a direct measure of information loss. Aim-level clinical validity rests not on this analysis but on the independently observed outcomes reported above.

## Conclusion

The early intensive care course of acute stroke can be represented as movement among a small number of interpretable dynamic states. This representation transported to an independent multicentre cohort, preserving its profiles, dynamics, outcome associations and prospective information, with limited between-hospital clustering. It was not, however, independently rediscovered in full: neither de novo solution recovered the neurological impairment-low support state, while the other three were matched closely by both. Transportability and independent rediscovery are distinct properties of a latent-state model and should be reported separately.

## Data Availability

MIMIC-IV v3.1 and the eICU Collaborative Research Database v2.0 are held at PhysioNet under credentialed access and are governed by the PhysioNet Credentialed Health Data Use Agreement 1.5.0. Neither may be redistributed by the authors. Access is available to any investigator who becomes a credentialed PhysioNet user, completes the CITI "Data or Specimens Only Research" training and signs the agreement. Patient-level intermediate files derived from these databases are governed by the same agreement and are therefore not deposited openly. The frozen model parameters are available from the corresponding author to any investigator holding credentialed access to MIMIC-IV. All analysis code is publicly available at https://github.com/ping-ai685/stroke-states-eicu

https://physionet.org/content/mimiciv/

https://physionet.org/content/eicu-crd/

## Data availability

The data underlying this article are the MIMIC-IV database, version 3.1, and the eICU Collaborative Research Database, version 2.0. Both are held at PhysioNet under credentialed access and are governed by the PhysioNet Credentialed Health Data License 1.5.0 and the corresponding Data Use Agreement 1.5.0. Neither may be redistributed by the authors. Access is available to any investigator who becomes a credentialed PhysioNet user, completes the CITI “Data or Specimens Only Research” training and signs the agreement, after which both databases can be obtained in full from https://physionet.org/content/mimiciv/ and https://physionet.org/content/eicu-crd/.

The same restriction extends to material derived from them. The patient-level intermediate files generated for this study — time-window matrices, state assignments and the prediction datasets — are governed by the agreement and are therefore not deposited in an open repository. The frozen parameters that define the transported model (emission and transition parameters, imputation and scaling constants, and state labels) are aggregate quantities containing no patient-level records, but they are derived from credentialed data and are therefore not posted openly; they are available from the corresponding author to any investigator who holds credentialed access to MIMIC-IV, so that the transport can be reproduced exactly rather than refitted as an approximation.

All analysis code contains no patient data and is publicly available at https://github.com/ping-ai685/stroke-states-eicu. It includes the scripts that export and verify the frozen discovery parameters, construct the eICU phenotype, cohort and time windows, apply the frozen model, fit the de novo and prediction models, and generate every table and figure reported here.

## Acknowledgements

**Use of generative AI.** The authors used generative AI assistants in preparing this work and disclose that use here, in accordance with the journal’s policy and the COPE position statement on authorship and AI tools.

Claude (Anthropic) was used to draft and edit the analysis code, to execute analyses at the authors’ direction, to generate tables and figures from the analysis output, to write the verification scripts that check every number and every stated claim in this manuscript against the analysis outputs, and to draft and revise manuscript text, including passages that interpret the findings. ChatGPT (OpenAI) was used to read manuscript drafts and return editorial comments; it was not used for code, study design or analysis.

The research question, the study design, the analysis protocol and its prespecified criteria, and the clinical adjudication of the stroke phenotype and the free-text term dictionaries are the authors’ own. Every AI-assisted output — code, results, figures and text — was reviewed by the authors, who directed the work throughout. The authors take full responsibility for the content of this article, including any part produced with AI assistance. Neither tool is listed as an author and neither meets authorship criteria.

## Ethics

This is a retrospective analysis of two de-identified databases distributed through PhysioNet under credentialed access: the eICU Collaborative Research Database v2.0[2] and MIMIC-IV v3.1.[3] Collection of the MIMIC-IV data and creation of that research resource were reviewed by the Institutional Review Board of Beth Israel Deaconess Medical Center, which granted a waiver of informed consent and approved the data sharing initiative. eICU-CRD is de-identified to the Safe Harbor provision of the US Health Insurance Portability and Accountability Act, certified by an independent third party (Privacert; HIPAA certification no. 1031219-2). The corresponding author completed the CITI Program course Data or Specimens Only Research and signed the PhysioNet Credentialed Health Data Use Agreement (v1.5.0) before accessing either database. As the work involved only secondary analysis of existing de-identified public datasets, no additional institutional review board approval or informed consent was required.

## Author contributions

PL conceived the study, wrote the analysis protocol, directed and verified all analyses, interpreted the results and drafted the manuscript. YX and YZ adjudicated the stroke phenotype and the free-text term dictionaries, reviewed the manuscript critically for important intellectual content, and contributed to the interpretation of the findings. All authors approved the final manuscript and agree to be accountable for all aspects of the work.

---

**Figure 1.**
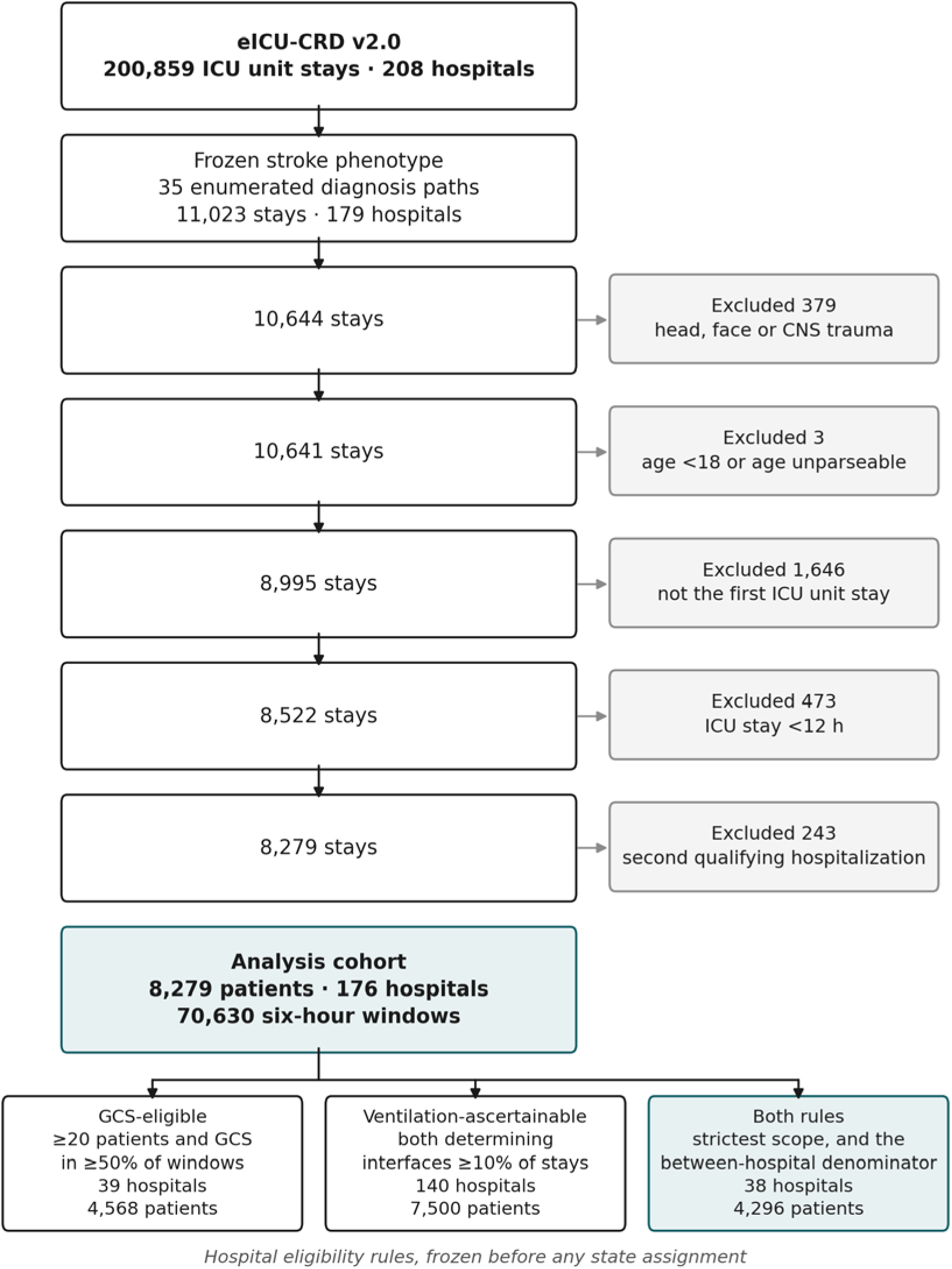
Cohort construction and hospital eligibility. Flow from eICU-CRD v2.0 to the analysis cohort of 8,279 patients across 176 hospitals. The stroke phenotype is an enumerated list of 35 diagnosisstring paths, frozen before extraction. Both hospital eligibility rules were frozen on counts and missingness alone, before any state was assigned, and define the analysis scopes used throughout. Alt text: A vertical flow diagram. The top box holds all eICU-CRD v2.0 stays matching the frozen 35-path stroke phenotype; five boxes below it show the cohort shrinking at each exclusion, with the number removed shown to the right of each step, ending at 8,279 patients in 176 hospitals. A branch at the foot shows the two hospital eligibility rules and the four analysis scopes they define.

**Figure 2.**
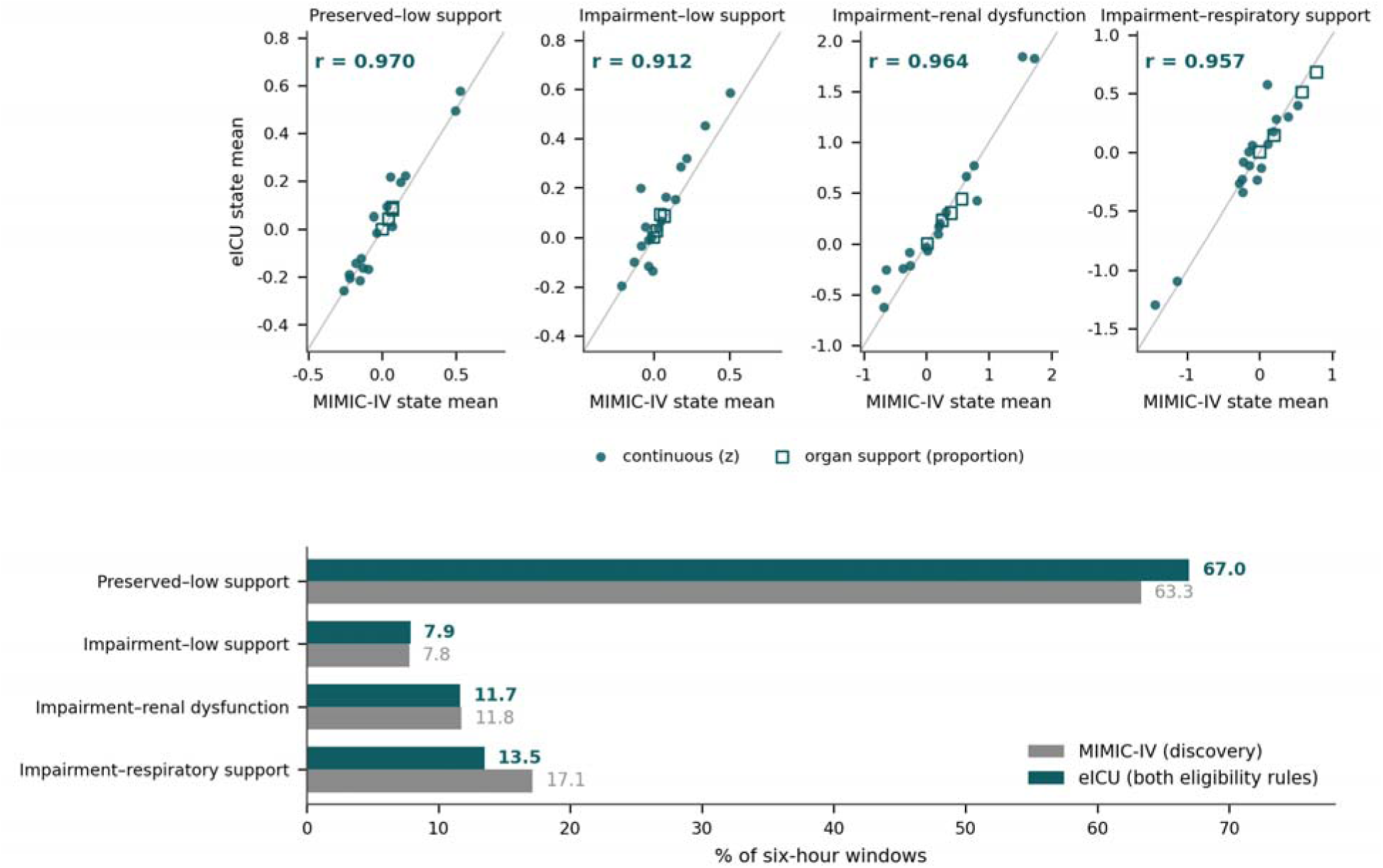
Frozen-model transport. *Upper row:* agreement between the MIMIC-IV and eICU state means for each of the 21 model variables, one panel per state, in the strictest analysis scope (hospitals meeting both eligibility rules). Filled circles are the 17 continuous variables on the frozen z-scale; open squares are the four organ-support variables as proportions. The grey line is identity; r is the profile correlation reported in Table 2. *Lower panel:* state prevalence as a percentage of six-hour windows, eICU against the discovery cohort. Alt text: Five panels. The four panels along the top are scatter plots, one per state, each plotting the eICU mean of a model variable against the MIMIC-IV mean, with a diagonal identity line; the points lie close to that line in every panel, and the correlation is printed in each. The panel below is a horizontal bar chart comparing the percentage of six-hour windows in each state between the two databases; the bars are of similar length for all four states.

**Figure 3.**
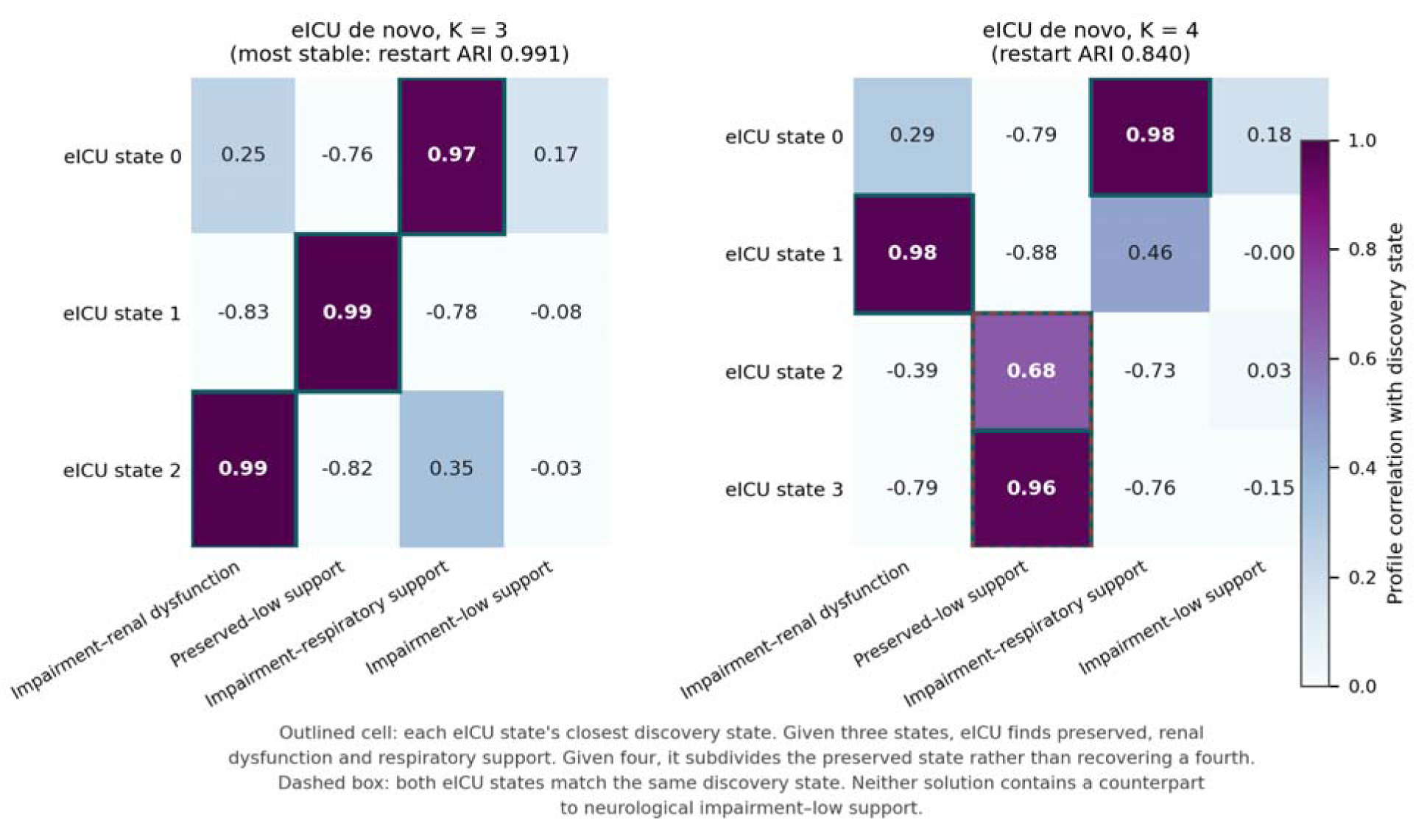
Structural reproducibility of the eICU de novo solutions. Profile correlation between each independently fitted eICU state and each discovery state, computed on feature profiles with no outcome information. *Left:* the three-state solution, which is eICU’s most stable (restart adjusted Rand index 0.991). *Right:* the four-state solution (0.840). Outlined cells mark each eICU state’s closest discovery state. The dashed box marks the two four-state solution states whose closest match is the same discovery state: given a fourth state, eICU subdivides the preserved state rather than recovering neurological impairment–low support, which has no counterpart in either solution. Alt text: Two heat maps side by side, the three-state solution on the left and the four-state solution on the right. Rows are the states eICU recovered on its own, columns are the four discovery states, and cell shading is the correlation between their feature profiles. Each row has one strongly shaded cell except in the column for neurological impairment–low support, which is pale throughout both maps. A dashed box on the right map marks two rows whose strongest cell falls in the same column.

**Figure 4.**
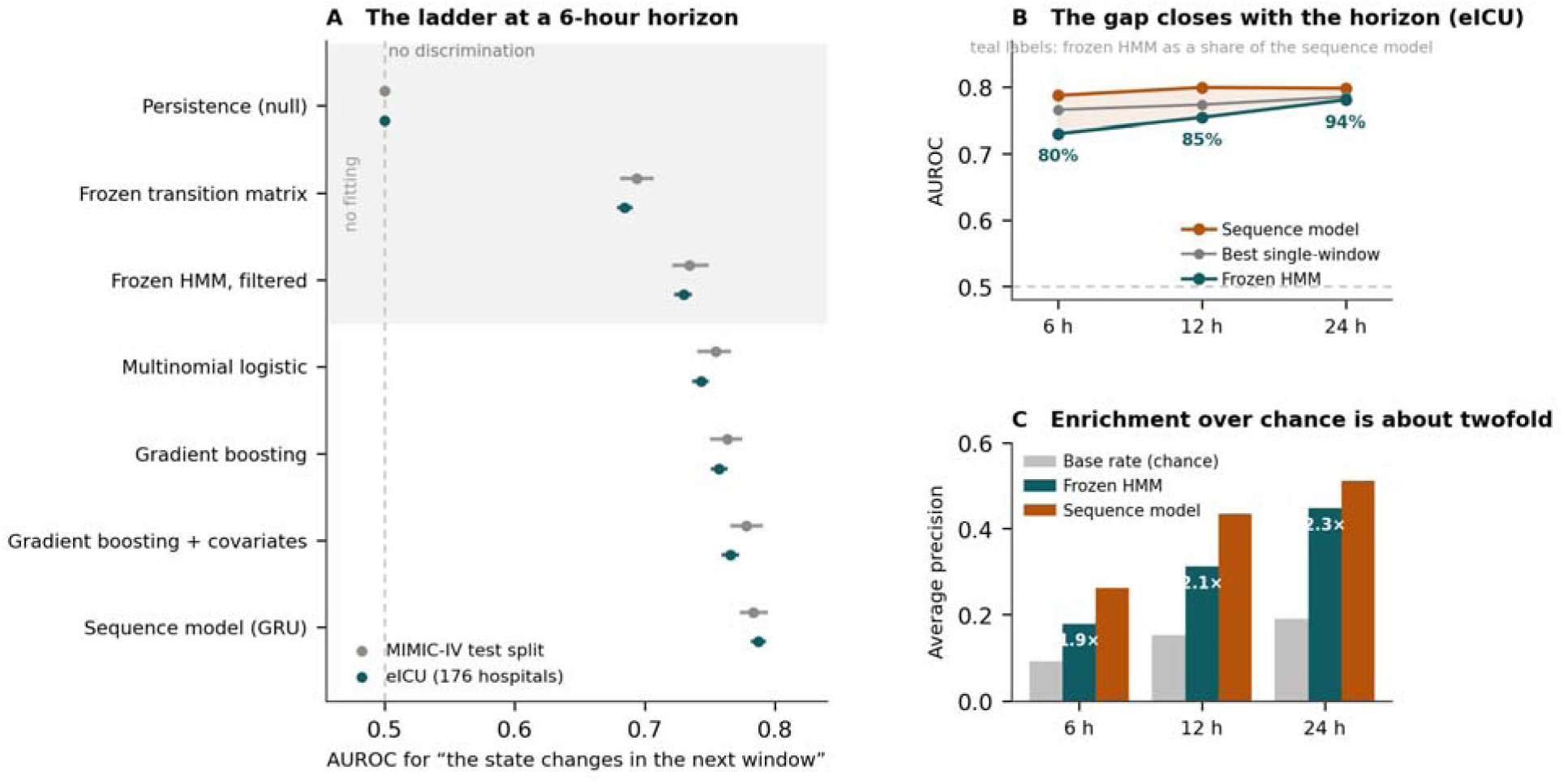
One-step-ahead prediction of the next state. *(A)* Discrimination for the event that the state differs in the following 6-hour window, for each predictor in the information ladder, in the held-out MIMIC-IV test split (grey) and in eICU (teal). Points are areas under the ROC curve and bars are patient-clustered bootstrap 95% intervals. Persistence sits at 0.5 by construction: it assigns the same probability to every window. The shaded band marks the three predictors that involve no fitting of any kind — they are the discovery model used for a different operation. Internal and external values sit close together at every rung. *(B)* The same comparison in eICU at three horizons, with the frozen transition matrix raised to the corresponding power and the fitted predictors trained separately in MIMIC-IV for each horizon and then frozen for external evaluation. Discrimination rises rather than decays, and the frozen representation rises far faster than the sequence model, so the two converge; the shaded area is the discrimination gap between the frozen representation and the sequence model, and it narrows as the horizon lengthens. Teal labels give the frozen model’s discrimination above the null as a percentage of the sequence model’s. *(C)* Average precision against the base rate of the event. Enrichment over chance is about twofold throughout. Together with a sensitivity of 27.8% at 90% specificity for the frozen model at six hours, this is why the analysis is reported as a property of the representation and not as a usable alarm. Alt text: Three panels. Panel A is a dot-and-interval plot of area under the ROC curve for seven predictors ordered by the information each uses, shown for MIMIC-IV and for eICU; values rise from 0.5 at the leftmost predictor to about 0.79 at the rightmost, and the two databases track each other closely. Panel B is a line plot of the same measure at 6, 12 and 24 hours for the frozen representation and the sequence model; both lines rise, the frozen line rises more steeply, and the shaded area between them narrows from left to right. Panel C plots average precision against the event base rate at each horizon, with every point roughly twice the base rate.

## References

[1] Lei P, Xu Y, Zhang Y. Dynamic clinical states and transitions during the first 72 hours of intensive care after acute stroke. medRxiv. Preprint posted online September 1, 2026. doi:10.64898/2026.08.30.26361738

[2] Pollard TJ, Johnson AEW, Raffa JD, Celi LA, Mark RG, Badawi O. The eICU Collaborative Research Database, a freely available multi-center database for critical care research. Sci Data. 2018;5:180178. doi:10.1038/sdata.2018.178

[3] Johnson AEW, Bulgarelli L, Shen L, et al. MIMIC-IV, a freely accessible electronic health record dataset. Sci Data. 2023;10(1):1. doi:10.1038/s41597-022-01899-x

[4] Goldberger AL, Amaral LAN, Glass L, et al. PhysioBank, PhysioToolkit, and PhysioNet: components of a new research resource for complex physiologic signals. Circulation. 2000;101(23):e215–e220. doi:10.1161/01.CIR.101.23.e215

[5] Schreiber J. pomegranate: fast and flexible probabilistic modeling in Python. J Mach Learn Res. 2018;18(164):1–6.

[6] Liang KY, Zeger SL. Longitudinal data analysis using generalized linear models. Biometrika. 1986;73(1):13–22. doi:10.1093/biomet/73.1.13

[7] Schwarz G. Estimating the dimension of a model. Ann Stat. 1978;6(2):461–464. doi:10.1214/aos/1176344136

[8] Hubert L, Arabie P. Comparing partitions. J Classif. 1985;2(1):193–218. doi:10.1007/BF01908075

[9] Pedregosa F, Varoquaux G, Gramfort A, et al. Scikit-learn: machine learning in Python. J Mach Learn Res. 2011;12:2825–2830.

[10] Cho K, van Merrienboer B, Gulcehre C, et al. Learning phrase representations using RNN encoder-decoder for statistical machine translation. In: Proceedings of the 2014 Conference on Empirical Methods in Natural Language Processing (EMNLP). Association for Computational Linguistics; 2014:1724–1734. doi:10.3115/v1/D14-1179

